# The role of pregnancy- related comorbidities in maternal health disparities among Asian American/Pacific Islanders

**DOI:** 10.64898/2026.07.10.26357771

**Authors:** Mekhala V. Dissanayake, Maya B. Mathur, Elliott K. Main, Suzan L. Carmichael

**Affiliations:** Department of Pediatrics, Stanford University School of Medicine, Palo Alto, CA; Quantitative Sciences Unit, Stanford University School of Medicine, Palo Alto, CA; Department of Obstetrics and Gynecology, Stanford University School of Medicine, Palo Alto, CA

## Abstract

**Background:** Asian American/ Pacific Islander (AAPI) groups experience different burdens of severe maternal morbidity (SMM) and pregnancy-related comorbidities that contribute to SMM. We sought to estimate counterfactual disparities for selected AAPI groups, separating out the pathway that includes these comorbidities.

**Methods:** We used linked birth certificates and hospital discharge data from births in California from 2011-2020. We examined the presence of pregnancy-related comorbidities (gestational hypertension and diabetes, pre-eclampsia) as a mediator between race/ethnicity and severe maternal morbidity. We used marginal structural models to estimate total effects and controlled direct effects (CDEs) (adjusted for maternal characteristics) for each AAPI group with Non-Hispanic Whites as the referent group.

**Results:** Our sample included n=1,849,698 births. All AAPI groups had higher prevalences of comorbidities compared to Non-Hispanic Whites (e.g., White: 15.7%, Chinese: 17.9%: Indian: 25.2%, Filipina: 28.8%, Pacific Islander: 23.9%). Filipinas and Pacific Islanders experienced the largest disparities in SMM (e.g., Total effect risk ratios (RR) Chinese: 1.03 (95% Confidence Interval (CI): 1.00, 1.07); Filipina: 1.64 (95% CI: 1.58, 1.70)). Under M=1 conditions, where everyone experienced comorbidities, disparities were eliminated for Chinese and Indian groups and alleviated for Filipinas and Pacific Islanders (e.g., CDE Chinese: 0.75 (95% CI: 0.69, 0.81); Filipina: 1.21 (95% CI: 1.13, 1.29)). Under M=0, disparities remained similar to the total effect (e.g., CDE Chinese: 1.14 (95% CI: 1.09, 1.19); CDE Filipina: 1.64 (95% CI: 1.53, 1.71))

**Conclusions:** Pregnancy-related comorbidities contributed substantially to disparities for AAPI groups. Disparities persisted for Filipinas and Pacific Islanders, suggesting a need for tailored interventions.

## Introduction

The health outcomes of Asian American/ Pacific Islanders (AAPI) groups are often aggregated, despite these groups’ diverse backgrounds, immigration histories, and social standings in the United States.^1,2^ In addition to having different histories of colonization, some AAPI groups are more likely to have had many generations living in the U.S. while other groups comprise recent immigrants who may be refugees or workers on specific visas.^1–3^ These different trajectories likely contribute to considerable variation with respect to income, discrimination, and acculturation, all of which may affect health outcomes.^1,3^ Therefore, aggregating health outcomes likely masks disparities for some.^1,2^

For example, in the case of severe maternal morbidity – the unintended consequences of labor and delivery with severe health implications – we previously estimated an SMM prevalence of 1.10% among all AAPI women birthing in California.^4,5^ When disaggregated, we found prevalences of 1.7% for Filipina women, 1.5% for Thai women, 1.3% for Hmong women, and 1.3% for Indian women.^6^ Compared to Non-Hispanic White women, these disparities approach what we observed for Non-Hispanic Black women in California.^4^

Previous research has found that this pattern of differences persists across perinatal outcomes and pregnancy-related comorbidities.^7–10^ Both hypertensive disorders of pregnancy (gestational hypertension and preeclampsia) and gestational diabetes are associated with increased risk of SMM.^11–14^ Pregnancy associated hypertension is on the rise in the U.S., including among all Asian subgroups, and in particular among Filipinas who have had the highest burden.^15^ Multiple studies across U.S. populations and data sources have reported that Indian, Filipina, Samoan, Guamanian, and Hawaiian women have the highest prevalence of hypertensive disorders of pregnancy, in additional to substantial variation in perinatal outcomes.^7,15–18^ Studies that disaggregate gestational diabetes risk found that Vietnamese, Filipina, Chinese, and Indian women have the highest burdens.^7,19^

Given the heterogeneity of these groups, it is likely that the drivers of SMM disparities are similarly heterogeneous. However, previous research has not determined how drivers of SMM disparities may vary between different AAPI groups. This analysis sought to fill this research gap by estimating counterfactual disparities, separating out the pathway that includes these comorbidities, with the goal of informing potentially different strategies for intervention for AAPI groups. We conducted our analysis in the state of California, which has a diverse AAPI population and the most births to AAPI people in the U.S.^20^

## Methods

### Dataset/ Population

This was a population-based analysis of data from the California Department of Health Care Access and Information (HCAI). Almost 100% of births in California are included in this resource (excluding home births and other unlinkable births).^21^ In this study, we used vital statistics of live birth and fetal death files linked to maternal hospital discharge records, ambulatory files, and emergency room admissions up to 42 days postpartum. We restricted our sample to births (both live and stillbirth) occurring between 20-45 weeks of gestation from 2011-2020 and retained one record each multiple birth (e.g., twins). We limited our analytic sample to individuals reporting non-Hispanic White, Indian, Chinese, Filipina, or Pacific Islander as their race/ethnicity (individuals reporting more than one race/ethnicity or Hispanic ethnicity were excluded). These Non-Hispanic AAPI groups were selected based on previous literature suggesting elevated risk of SMM, elevated risk of the comorbidities under study, and sufficient sample size to conduct mediation analyses.^1,6,7,15–18^

### Outcome

Our outcome was SMM at delivery and up to 42 days postpartum, captured in the linked maternal hospital discharge records (delivery and postpartum), ambulatory files (postpartum), and emergency room admissions (postpartum). We used the updated SMM Index, developed by the United States Centers for Disease Control and Prevention, to identify SMM events.^22^ This composite includes 21 indicators that are identified using International Classification of Diseases, 9^th^ and 10^th^-Revision (ICD-9/ICD-10) diagnosis and procedure codes. Birthing people with one or more of these indicators occurring either during delivery or in the postpartum period were considered to have SMM.

### Exposure

Our exposure for the mediation analysis was race/ethnicity, determined from the vital statistics records. We define race not as a biological trait, but as a social construct whose meaning is reified in social structures and is a marker of experiences and exposures.^23^ We conducted mediation analyses for Chinese, Indian, Filipina, and Pacific-Islander groups. Non-Hispanic Whites served as the reference group for our mediation analyses, because they are the majority and dominant racial/ethnic group in the United States, and our goal was to quantify disparities.^24^

### Mediator

Estimating path-specific effects – meaning causal effects of a single mediator – would require knowledge of the timing of different mediators that was not available in our data. We therefore examined joint causal effects of our mediators.^25^ Our mediators were pregnancy-related comorbidities, including gestational hypertension, pre-eclampsia, and gestational diabetes. These were identified by either 1) checkboxes on the vital statistics records or 2) ICD codes in the linked hospital discharge record from the delivery.

### Covariates

We used a Directed Acyclic Graph (DAG) to choose covariates for our analysis (Figure 1). We included the following, also derived from either the vital statistics records or the linked hospital discharge record from the delivery: Maternal age (<20, 20-24, 25-29, 30-34, 35-39, 40+ years), Parity (Nulliparous/Multiparous), pre-pregnancy Body Mass Index (BMI) (Underweight <18.5 kg/m^2^, Normal Weight 18.5-<24.9 kg/m^2^, Overweight 25-<29,9 kg/m^2^, Obese 30+ kg/m^2^ kg/m^2^), Education (Less than high school, High school grad, Some college, Undergraduate, Postgraduate), and Insurance, defined as the payer from the delivery hospitalization (Private, Public, Uninsured/Self-Pay, Other).

**Figure 1:**
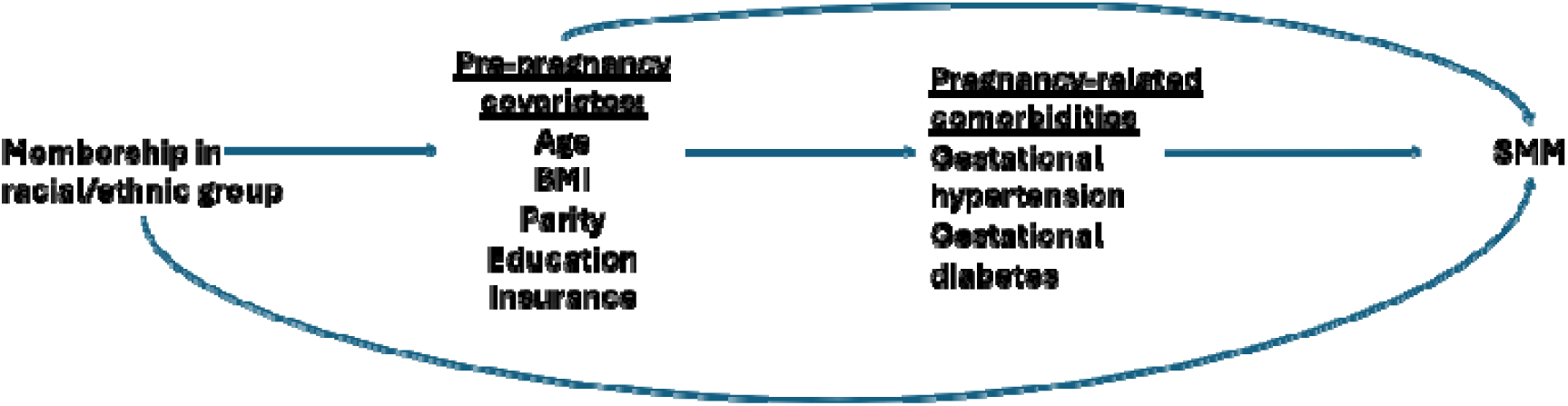
A simplified Directed Acyclic Graph (DAG) depicting our understanding of relationships between variables in our study. We conceptualize race/ethnicity not as a biological variable, but rather the social category of membership in a racial/ethnic group that leads to disparities in our outcome of SMM. There are other historical and contemporary processes that are not depicted in this diagram. As depicted here, pregnancy-related covariates are confounders from the perspective of the mediator-outcome relationship, but mediators of the exposure-outcome relationship.

### Statistical Analysis

We first conducted a descriptive analysis of all variables by racial/ethnic group. We conducted mediation analyses for each AAPI group separately, contrasting each group with our White reference group (e.g., Indian/White, Chinese/White, etc.). We used marginal structural models to conduct our mediation analyses because we were interested in isolating the controlled direct effect (CDE), or the effect of race on SMM that does not involve the pathway with pregnancy-related comorbidities.^24,25^ We used marginal structural models to estimate average counterfactual outcomes and account for time-varying confounding, present in our analysis because pregnancy-related covariates were confounders from the perspective of the mediator-outcome relationship, and mediators of the exposure-outcome relationship (Figure 1).^25^ The CDE in this context is a disparity measure, estimating what the racial disparity in SMM would be if mediators were set to the same level in the population.^24,25^

Because all measured covariates were temporally downstream of maternal racial/ethnic identity, we did not include weights for exposure-outcome confounding. To obtain the total effect, we regressed the outcome, SMM, on race/ethnicity using Poisson models to estimate the risk ratio. To account for mediator-outcome confounding in our mediation models, we fit inverse probability weights using logistic regression models. To obtain the CDE, we regressed SMM on race/ethnicity (main effect), pregnancy-related comorbidities (mediator), and an interaction between the two, and included the constructed weights. We bootstrapped analyses (1000 replicates) and used the percentile method to obtain 95% confidence intervals. From these parameters, we calculated the CDE under two conditions – when the mediator was held at a constant value of 1 (i.e., the counterfactual disparity when the entire population had a pregnancy-related comorbidity) or 0 (i.e., the counterfactual disparity when no one had a pregnancy-related comorbidity). We interpreted the CDEs as the remaining, or unexplained, disparities if pregnancy-related comorbidities were set to the same value across racial groups.

We conducted three sensitivity analyses. First, we repeated all analyses with a revised SMM definition, removing cases where transfusion was the only indicator of SMM (‘non-transfusion SMM’).^26^ Second, because previous research has conceptualized nativity as an effect modifier,^1^ we stratified analyses by foreign-born status by conducting analyses separately for US-born AAPI groups and foreign-born AAPI groups, with all White individuals still as the referent group. Lastly, we calculated mediational E-values to determine whether unmeasured confounding could explain our findings.^27^ This study was approved by the Committee for the Protection of Human Subjects (17–04–2932) and [redacted for review] IRB (43029) protocols. We conducted analyses in SAS version 9.4 and R version 2025.05.1.

## Results

Table 1 summarizes characteristics of our study population by race/ethnicity (n=1,849,698 births). Obesity was less common among all AAPI groups compared to Whites, with the notable exception of Pacific Islanders (White: 18.3%; Pacific Islander: 44.2%). Births to Chinese and Indian groups were more likely to have postgraduate degrees and less likely to have public insurance than Whites, while the reverse was true for Pacific Islanders. The lowest percentage of US-born births occurred among Indians (9.3%) and the highest among Pacific Islanders (65.7%).

**Table 1:** Descriptive characteristics of study population by race/ethnicity^a^, births in California 2011-2020 (N=1,849,698)

|  |  |  |  |  |  |
| --- | --- | --- | --- | --- | --- |
|  | White<br>(n= 1,148,243) | Chinese<br>(n= 183,548) | Indian<br>(n= 109,353) | Filipina<br>(n= 108,642) | Pacific Islander<br>(n= 24,055) |
| Gestational Hypertension/<br>Preeclampsia | 103,334 (9.0) | 7,377 (4.0) | 7,665 (7.0) | 13,915 (12.8) | 3,003 (12.5) |
| Gestational Diabetes | 91,897 (8.0) | 27, 428 (14.9) | 22,029 (20.1) | 21,548 (19.8) | 3,523 (14.7) |
| Any pregnancy-related<br>comorbidity | 180,058 (15.7) | 32,871 (17.9) | 27,597 (25.2) | 31,315 (28.8) | 5,755 (23.9) |
| **Maternal Age** |  |  |  |  |  |
| <20 | 24306 (2.1) | 152 (0.1) | 161 (0.2) | 1,153 (1.1) | 1,152 (4.8) |
| 20-24 | 134,336 (11.7) | 4,632 (2.5) | 4,648 (4.3) | 7,979 (7.3) | 5,315 (22.1) |
| 25-29 | 296,045 (25.8) | 37,487 (20.4) | 30,199 (27.6) | 23,046 (21.2) | 7,190 (29.9) |
| 30-34 | 405,800 (35.3) | 76,442 (41.7) | 50,668 (46.3) | 39,335 (36.22) | 6,345 (26.4) |
| 35-39 | 231,342 (20.2) | 52,068 (28.4) | 20,626 (18.9) | 29,175 (26.9) | 3,237 (13.5) |
| 40+ | 56,414 (4.9) | 12,767 (7.0) | 3,051 (2.8) | 7,954 (7.3) | 816 (3.4) |
| **Nulliparous** | 500,518 (43.6) | 91,873 (50.1) | 54,845 (50.2) | 45,058 (41.5) | 8,840 (36.8) |
| **Pre-pregnancy BMI** |  |  |  |  |  |
| Underweight | 43,753 (3.8) | 23,720 (12.9) | 4,949 (4.5) | 4,271 (3.9) | 459 (1.9) |
| Normal Weight | 626,469 (54.6) | 133,285 (72.6) | 64,474 (59.0) | 61,009 (56.2) | 6,806 (28.3) |
| Overweight | 267,653 (23.3) | 21,768 (11.9) | 29,953 (27.4) | 28,618 (26.3) | 6,158 (25.6) |
| Obese | 210,368 (18.3) | 4,775 (2.6) | 9,977 (9.1) | 14,744 (13.6) | 10,632 (44.2) |
| **Education** |  |  |  |  |  |
| Less than high school | 45,167 (3.9) | 2,576 (1.40) | 1,935 (1.8) | 1,322 (1.2) | 1,842 (7.7) |
| High school grad | 204,913 (17.9) | 13,107 (7.1) | 7,696 (7.0) | 11,104 (10.2) | 8,929 (37.1) |
| Some College | 320, 641 (27.9) | 27,081 (14.8) | 9,226 (8.4) | 35,834 (33.0) | 9,143 (38.0) |
| Undergraduate | 359,976 (31.4) | 77,563 (42.3) | 38,790 (35.5) | 49,319 (45.4) | 3,142 (13.1) |
| Postgraduate degree | 217,546 (19.0) | 63,221 (34.4) | 51,706 (47.3) | 11,063 (10.2) | 999 (4.2) |
| **Insurance** |  |  |  |  |  |
| Private | 841,663 (73.3) | 103,069 (56.2) | 89,535 (81.9) | 82,758 (76.2) | 12,425 (51.7) |
| Public | 284,365 (24.8) | 21,442 (11.7) | 15,372 (14.1) | 23,156 (21.3) | 11,112 (46.2) |
| Uninsured/ Self-Pay | 12,256 (1.1) | 57,132 (31.1) | 656 (0.6) | 831 (0.8) | 239 (1.0) |
| Other | 9,959 (0.9) | 1,905 (1.0) | 3,790 (3.5) | 1,897 (1.8) | 279 (1.2) |
| **Nativity** |  |  |  |  |  |
| US Born | 982,996 (85.6) | 26,073 (14.2) | 10,190 (9.3) | 35,220 (32.5) | 14,738 (65.7) |
| **SMM** |  |  |  |  |  |
| Overall SMM | 18,768 (1.6) | 3,149 (1.7) | 2,058 (1.9) | 2,915 (2.7) | 642 (2.7) |
| Non-transfusion SMM<sup>b</sup> | 9,884 (0.9) | 1,447 (0.8) | 1,070 (1.0) | 1,518 (1.4) | 311 (1.3) |
| <sup>a</sup>All groups include only non-Hispanic individuals<br><sup>b</sup>Transfusion as only indicator excluded |  |  |  |  |  |

While Whites had a higher prevalence of gestational hypertension/ pre-eclampsia than Chinese or Indian groups, all AAPI groups had higher burdens of gestational diabetes. Following that pattern, all AAPI groups had higher prevalences of any pregnancy-related comorbidity. Filipina and Pacific Islanders had the highest burdens of both conditions (Filipina hypertension: 12.8%, diabetes 19.8 %; Pacific Islander hypertension: 12.5 %, diabetes 14.5%). The risk of SMM was highest among Filipinas and Pacific Islanders (2.7%).

Table 2 shows the estimates for all parameters from total effect and mediation models. The total effect models estimate existing disparities between each AAPI group and Whites. Filipina and Pacific Islander groups experienced the largest disparities in SMM, with risk ratios (RR) of 1.64 (95%CI: 1.58, 1.70) and 1.67 (95% CI: 1.55, 1.80), respectively. Given the similarity in patterns for all groups, we describe the results for Filipinas as an example. In the mediation model, the main effect for Filipina race/ethnicity was similar to the total effects model (1.62, 95% CI: 1.53, 1.71). The mediator effect of pregnancy-related comorbidities was much stronger, with a risk ratio of 2.53 (95% CI, 2.45, 2.62). However, there was a negative interaction between the effect of race/ethnicity and pregnancy-related comorbidities, suggesting that the effect of pregnancy-related comorbidities was attenuated among Filipinas (RR: 0.75, 95% CI: 0.69, 0.81).

**Table 2:** Risk ratios for AAPI groups^a^ from total effects and mediation models, California births 2011-2020.

|  | White<br>(n= 1,148, 243) | Chinese<br>(n= 183,548) | Indian<br>(n= 109,353) | Filipina<br>(n= 108,642) | Pacific Islander<br>(n= 24,055) |
| --- | --- | --- | --- | --- | --- |
| **Total effects model** |  |  |  |  |  |
| Main effect (race/ethnicity/ethnicity) | ref | 1.03 (1.00, 1.07) | 1.14 (1.10, 2.0) | 1.64 (1.58, 1.70) | 1.67(1.55, 1.80) |
| **Mediation model** |  |  |  |  |  |
| Main effect (race/ethnicity/ethnicity) |  | 1.14 (1.09, 1.19) | 1.24 (1.17, 1.31) | 1.62 (1.53, 1.71) | 1.64 (1.48, 1.82) |
| Mediator (pregnancy-related comorbidity) | ref | 2.54 (2.45, 2.62) | 2.53 (2.45, 2.62) | 2.53 (2.45, 2.62) | 2.52 (2.44, 2.6) |
| Interaction (race/ethnicity/ethnicity*comorbidity) | ref | 0.66 (0.60, 0.72) | 0.63 (0.56, 0.69) | 0.75 (0.69, 0.81) | 0.74 (0.61, 0.87) |

<sup>a</sup>All groups include only non-Hispanic individuals
Interpretation of risk ratios: Under the M=0 condition, the controlled direct effect (CDE) is given by the main effect from the mediation model (in the contrast between groups, the mediator and interaction terms fall out). Under the M=1 condition, the CDE is given by both the main effect and interaction term from the mediation model (in the contrast between groups, only the mediator term falls out).

Figure 2 displays total effects and CDEs under counterfactual conditions. For all groups under the hypothetical M=1 condition, because of the negative interaction, the CDE was lower than the total effect. For the Filipina and Pacific Islander groups, the CDE still indicated a disparity (RR>1) even under the M=1 hypothetical condition. However, among Chinese and Indian groups, whose total effects indicated a smaller overall disparity, the CDE effect indicated a disparity under the M=0 condition, but not when M=1. This suggests the Chinese and Indian groups may have *better* SMM outcomes than the White group if all individuals had pregnancy-related comorbidities (e.g. Chinese Total effect RR: 1.03, 95% CI: 1.0, 1.07), CDE RR: 0.75, 95% CI: 0.70, 80). For Filipina and Chinese groups, who experienced larger disparities, the CDE suggests disparities would be considerably reduced but not eliminated if these pregnancy-related comorbidities were held constant (e.g. Pacific Islander total effects RR: 1.67, 95% CI: 1.55, 1.80); CDE RR: 1.21, 95% CI: 1.05, 1.40). When M=0, however, for Chinese and Indian birthing people, the CDE was higher than the total effect (e.g., Chinese CDE: 1.14, 95% CI 1.09, 1.19). For Filipina and Pacific Islander groups, the CDE and total effect were similar (e.g. Pacific Islander CDE: 1.64 (95% CI: 1.48, 1.82). These results suggest the disparities in SMM are stronger in the hypothetical setting where no one has a pregnancy-related comorbidity.

**Figure 2:**
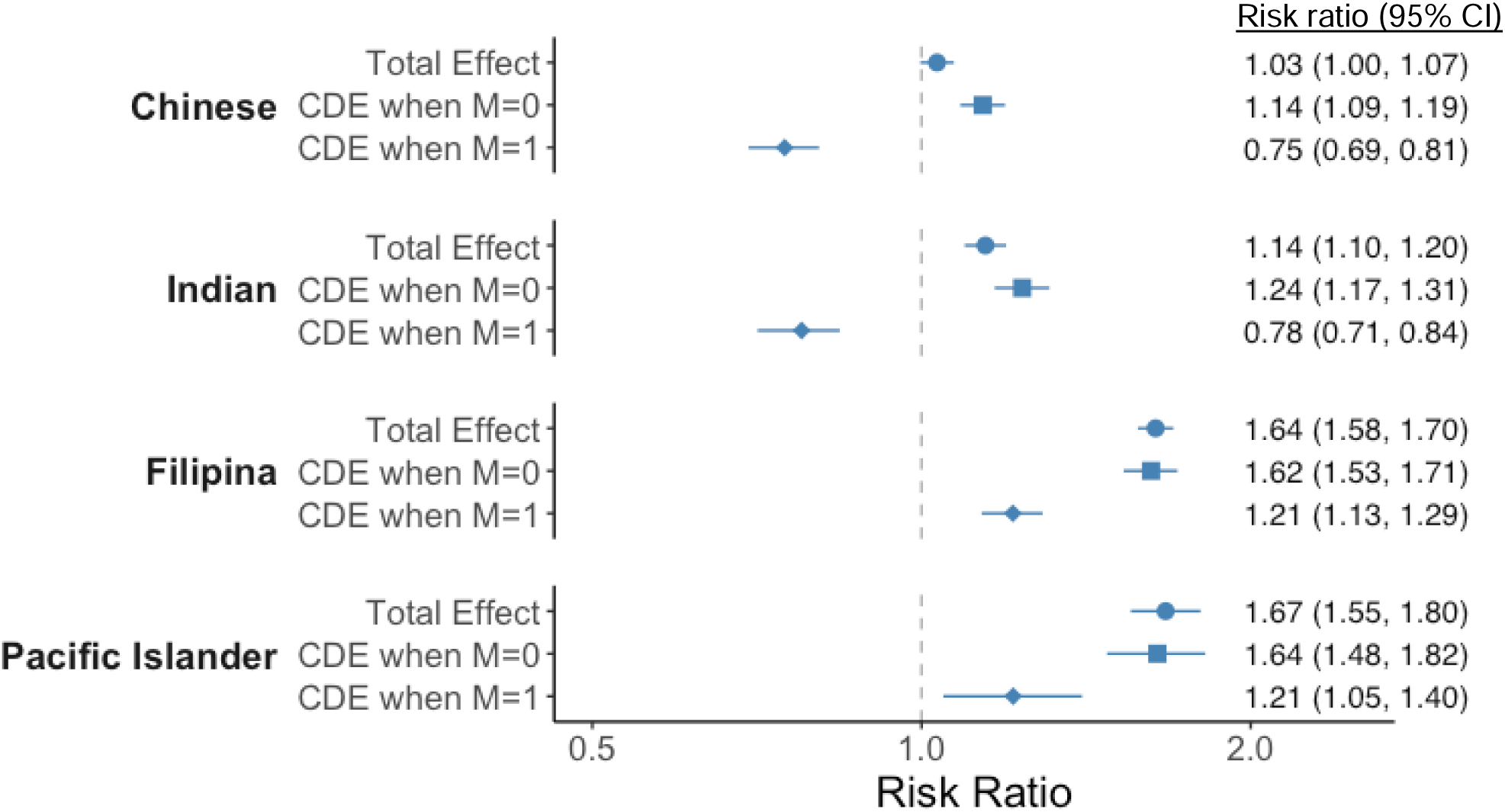
Forest plot depicting total effect and controlled direct effect (CDE) estimates for AAPI groups. For all risk ratios, the referent group is White. CDEs are presented for two hypothetical interventions: one preventing all pregnancy-related comorbidities (M=0) and one ensuring all individuals experienced pregnancy-related comorbidities (M=1).

In sensitivity analyses with non-transfusion SMM as the outcome, we observed similar patterns for all groups, though with larger confidence intervals associated with our estimates (Figure S1). We were unable to stratify all analyses by nativity given some sample size limitations (e.g., ∼10,000 US-Born Indian births, ∼14,000 US-Born Pacific Islander births). Descriptive characteristics for Filipina and Chinese groups stratified by nativity are available in the eAppendix (eTable 1). When stratifying by nativity, we found similar patterns between the total effect and CDEs (eFigure 2 and eFigure 3). However, among Filipina births, we found larger disparities in SMM among those who were foreign born compared to US born (Total effect US Born: RR: 1.38, 95% CI: 1.26,1.52), Foreign Born (RR:1.73, 95% CI: 1.63, 1.83)), but the reverse among Chinese births (Total effect US Born RR: 1.13 (95% CI: 1.00, 1.27); Foreign Born RR 0.85, 95% CI: 0.80, 0.90). Therefore, although nativity did not modify the effect of the mediator, it did modify overall SMM disparities. We also calculated mediational E-values for all CDE estimates, with values ranging from 1.54 – 2.66 (Table S2). For example, for Pacific Islanders under the condition M=0, to completely explain away the CDE of 1.64, an unmeasured confounder would have to have a risk ratio of at least 2.66 with both race/ethnicity and pregnancy-related comorbidities, above and beyond all measured covariates.^27^

## Discussion

We found that pregnancy-related comorbidities accounted for a large proportion, but not all, of the disparity between selected AAPI groups and Non-Hispanic Whites. Under the condition that everyone had comorbidities, disparities were eliminated among Chinese and Indian groups and attenuated among Filipinas and Pacific Islander groups. However, disparities either worsened or remained the same under the condition that no one had comorbidities, emphasizing the disparities these groups experience regardless of these comorbidities. Given the differences in both existing and remaining disparities between these groups, our results support continuing to disaggregate AAPI groups when examining disparities.

We found a negative interaction between all racial groups and pregnancy-related comorbidities, suggesting detrimental effects of these comorbidities were attenuated in these groups. Importantly however, although the interaction was negative, these AAPI groups still experienced higher prevalences of pregnancy-related comorbidities and SMM, indicating salient disparities.^28^ We emphasize that because race is a social construct, the causal effects we examined are the downstream consequences of membership in these racial/ethnic groups in the United States context, including the historical and contemporary conditions that lead to health disparities. In the context of this analysis, we estimated the impacts of having a particular socially constructed identity, not the effects changing that identity.^24,29^ We took a more pragmatic approach of interpreting counterfactual health disparities by focusing on a mediating factor downstream of race/ethnicity.

Similar work has been conducted on Black-White disparities in SMM. A study on the role of hypertensive disorders of pregnancy (HDP) in Black-White disparities found that about 26% of the Black-White rate difference in SMM would be eliminated if the pathway through HDP was blocked.^30^ With the caveat that we took different approaches to mediation and therefore our interpretations are also different, we found much larger reductions in disparities under the condition that no one had comorbidities, suggesting that drivers of disparities vary for different racial/ethnic groups.

We observed that some of the disparity in SMM remained for Filipina and Pacific Islander birthing people under the M=1 condition. These groups also had the highest burdens of SMM and pregnancy-related comorbidities. Other, unmeasured factors may also be important contributors to these disparities: studies conducted in the Mediators of Atherosclerosis in South Asians Living in America (MASALA) Cohort have recently looked at the contributions of immigration/acculturation ^31^ and discrimination, inflammation and coping^32^ to various health outcomes; these approaches could also be taken to understand conditions during pregnancy. Additionally, understanding social determinants of health, such as contributions of contextual factors like neighborhood-level resources, will enrich our understanding of drivers of disparities.^1^ For example, when examining SMM disparities at the intersection of race/ethnicity, weathering, neighborhood disenfranchisement and socioeconomic disadvantage, Pacific Islander individuals had higher predicted probabilities of SMM than their Asian counterparts, and further disaggregation of these groups may reveal further disparities.^33^

Strengths of this study include a large and diverse AAPI population, including SMM through the postpartum period, disaggregating AAPI groups, and aiming to determine drivers of disparities. One limitation is the mediation approach we used relies on strong assumptions of no confounding.^25^ Though residual or unmeasured mediator-outcome confounding remains possible, our mediational E-values suggest the strength of exposure-outcome and mediator-outcome relationships would have to be quite strong to fully explain our results. We note that the mediational E-values we calculated were originally tested for the natural direct or indirect effect, and with the CDE, we must make an additional assumption that if there is an unmeasured confounder “U” of the mediator-outcome relationship, that the exposure does not affect “U”.^27^

We only examined births to non-Hispanic, single-race/ethnicity individuals in this analysis, which limits our interpretation to these populations. An additional limitation is that we did not adjust for some chronic conditions, including chronic (pre-pregnancy) hypertension or diabetes. Chronic hypertension and diabetes are mutually exclusive of gestational hypertension and diabetes, because gestational hypertension and diabetes represent new onset of disease during pregnancy.^34^ Since there is only one checkbox for gestational hypertension and pre-eclampsia on the vital records, we could not separate out those with pre-eclampsia superimposed on chronic hypertension from those with who erroneously may have both gestational hypertension and chronic hypertension recorded, and therefore, we chose not to adjust for these conditions. However, given the relative strength of associations of our mediator, the bias from these conditions would have to be quite large to change our results, and chronic hypertension and diabetes are much less prevalent in our population than the pregnancy-related conditions.

Our approach does not quantify how management of these conditions may contribute to disparities. The World Health Organization has recommended lower BMI cutoffs for Asian populations for screening for type 2 diabetes and cardiovascular complications, given the association that exists at lower BMIs compared to other populations.^35^ Research on risk factors for gestational diabetes supports this, finding larger population attributable fractions when these cutoffs are used.^36^ Previous research has found that among all racial/ethnic groups studied, the progression of HDP to cardiovascular complications (including conditions that are part of SMM) was most common among AAPI birthing people.^37^ All of this considered, there may be additional nuances to screening and prevention of pregnancy-related comorbidities and the management of these conditions that contribute to disparities in SMM that we were unable to assess in our analysis.

By conducting mediation analyses among disaggregated AAPI groups in California, we found that the pregnancy-related comorbidities contributed significantly to disparities in SMM. Future research should investigate how both the prevention of these comorbidities and management of these conditions during pregnancy may vary for these groups and contribute to disparities in SMM. Further, research should also focus on other potential drivers of these disparities, including processes related to immigration, discrimination, and other social determinants of health.

## Supporting information

eTable1

eFigure1

eFigure2

eFigure3

eTable2

## Data Availability

Investigators trained in human subject research may request the data used in this study from the California Department of Health Care Access and Information.

## Notes

### Competing Interest Statement

The authors have declared no competing interest.

### Author Declarations

The Committee for the Protection of Human Subjects and IRB of Stanford University gave ethical approval for this work.

### Funding

- National Institute of Nursing Research: R01NR017020, R01NR017020
- Eunice Kennedy Shriver National Institute of Child Health and Human Development: U54 HD113142, R01HD103662

