## Supplementary material for "The role of pregnancy- related comorbidities in maternal health disparities among Asian American/Pacific Islanders": eTable1

| eTable 1: Descriptive characteristics of Chinese and Filipina groups stratified by nativity, California births, 2011-2020 | | | | |
| --- | --- | --- | --- | --- |
|  | Chinese | | Filipina | |
|  | US-born  (n= 26, 073) | Foreign Born  (n= 157,430) | US-born  (n= 31, 574) | Foreign Born  (n= 73,160) |
| Gestational Hypertension/ Preeclampsia | 2,064 (7.9) | 5,310 (3.4) | 4,637 (13.2) | 9,234 (12.6) |
| Gestational Diabetes | 4,158 (16.0) | 23,257 (14.8) | 5,945 (16.9) | 15,554 (21.3) |
| Any pregnancy-related comorbidity | 5,722 (22.0) | 27,134 (17.2) | 9,355 (26.6) | 21,878 (29.9) |
| **Maternal Age** |  |  |  |  |
| <20 | 49 (0.2) | 103 (0.1) | 681 (1.9) | 468 (0.6) |
| 20-24 | 377 (1.5) | 4255 (2.7) | 3,454 (9.8) | 4,509 (6.2) |
| 25-29 | 3,063 (11.8) | 34,418 (21.9) | 8,143 (23.1) | 14,847 (20.3) |
| 30-34 | 12,564 (48.2) | 63,853 (40.6) | 13,567 (38.5) | 25,689 (35.1) |
| 35-39 | 8,436 (32.4) | 43,620 (27.7) | 7,947 (22.6) | 21,160 (28.9) |
| 40+ | 1,584 (6.1) | 111,81 (7.1) | 1,428 (4.1) | 6,487 (8.9) |
| **Nulliparous** | 13,832 (53.1) | 78,022 49.6) | 15,882 45.1) | 29,075 39.7) |
| **Pre-pregnancy BMI** |  |  |  |  |
| Underweight | 1,329 (5.1) | 22,386 (14.2) | 799 (2.3) | 3,462 (4.7) |
| Normal Weight | 18,470 (70.8) | 11,4784 (72.9) | 174,87 (49.7) | 43,371 (59.3) |
| Overweight | 4,636 (17.8) | 17,125 (10.9) | 1,0015 (28.4) | 18,532 (25.3) |
| Obese | 1,638 (6.3) | 3,135 (2.0) | 6,919 (19.7) | 7,795 (10.7) |
| **Education** |  |  |  |  |
| Less than high school | 60 (0.2) | 2,515  (1.6) | 563 (1.6) | 757 (1.0) |
| High school grad | 613 (2.4) | 12,492 (7.9) | 3,511 (10.0) | 7,563 (10.3) |
| Some College | 2,060 (7.9) | 25,014 (15.9) | 11,864 (33.7) | 23,878 (32.6) |
| Undergraduate | 12,480 (47.9) | 65,056 (41.3) | 13,995 (39.7) | 35,204 (48.1) |
| Postgraduate degree | 10,860 (41.7) | 52,353 (33.3) | 5,287 (15.0) | 5,758 (7.9) |
| **Insurance** |  |  |  |  |
| Private | 24,541 (94.1) | 78,493 (50.0) | 28,940 (82.2) | 53,622 (73.3) |
| Public | 1,091 (4.2) | 203,49 (12.9) | 5,562 (15.8) | 17,536 (24.0) |
| Uninsured/ Self-Pay | 92 (0.4) | 57,033 (36.2) | 150 (0.4) | 673 (0.9) |
| Other | 349 (1.3) | 1555 (1.0) | 568 (1.62) | 1329 (1.8) |
| **SMM** |  |  |  |  |
| Overall SMM | 526 (2.0) | 2,623 (1.7) | 822 (2.3) | 2,082 (2.9) |
| Non-transfusion SMM | 247 (1.0) | 1,200 (0.8) | 411 (1.2) | 1,100 (1.5) |
