## Supplementary material for "The role of pregnancy- related comorbidities in maternal health disparities among Asian American/Pacific Islanders": eFigure1

eFigure 1: Mediation results with transfusion-only SMM excluded

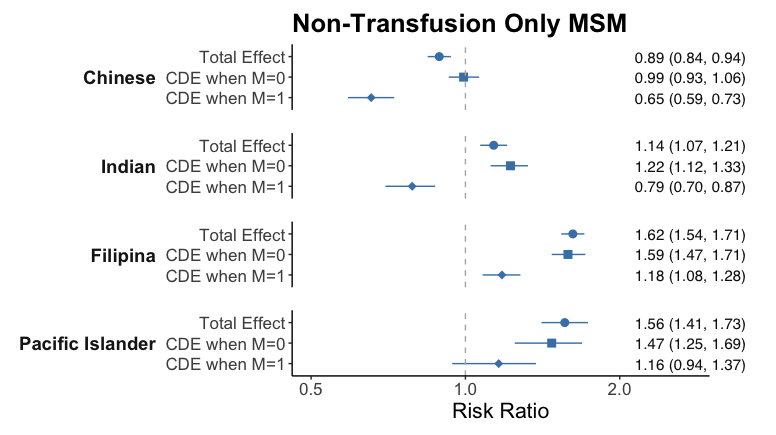

Risk ratio (95% CI)

Figure S2: Chinese Mediation results stratified by nativity
