## Supplementary material for "The role of pregnancy- related comorbidities in maternal health disparities among Asian American/Pacific Islanders": eFigure2

eFigure 2: Mediation results for Chinese births stratified by nativity

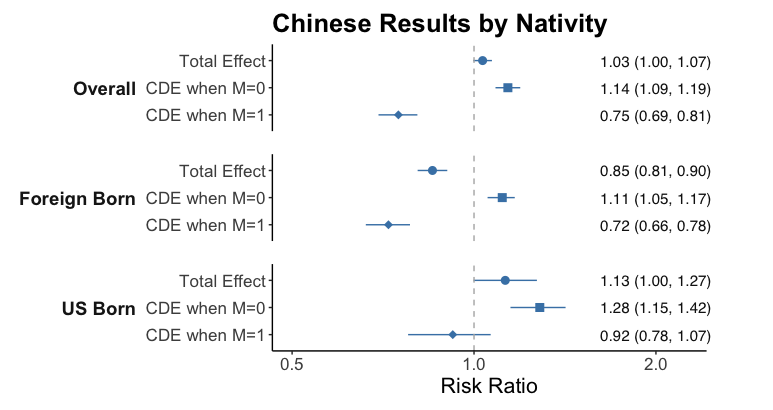

Risk ratio (95% CI)

Figure S3: Filipina mediation results stratified by nativity
