## Supplementary figures and images for "The role of pregnancy- related comorbidities in maternal health disparities among Asian American/Pacific Islanders"

### eFigure3

eFigure 3: Mediation results for Filipina births stratified by nativity

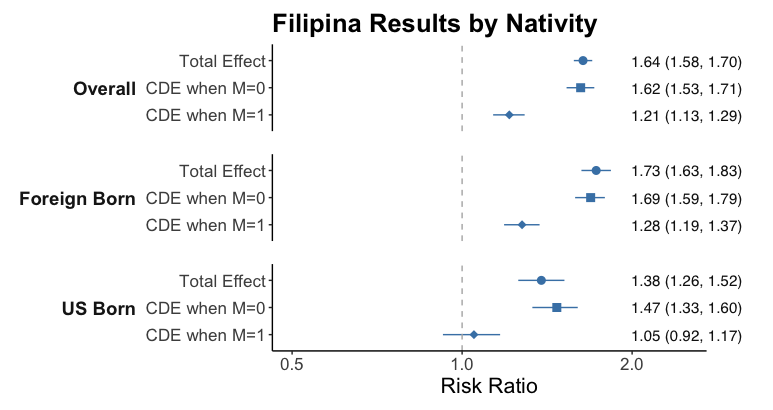

Risk ratio (95% CI)
