## Supplementary material for "The role of pregnancy- related comorbidities in maternal health disparities among Asian American/Pacific Islanders": eTable2

| eTable 2: Mediational E-values for Controlled Direct Effects (CDE) | | | |
| --- | --- | --- | --- |
| Race/Ethnicity | CDE Condition | CDE estimate | Mediational E-Value |
| Chinese | M=0 | 1.14 | 1.54 |
|  | M=1 | 0.75 | 2.00 |
| Indian | M=0 | 1.24 | 1.79 |
|  | M=1 | 0.78 | 1.88 |
| Filipina | M=0 | 1.62 | 2.62 |
|  | M=1 | 1.21 | 1.71 |
| Pacific Islander | M=0 | 1.64 | 2.66 |
|  | M=1 | 1.21 | 1.71 |
